# Mortality and health losses from cardiovascular diseases attributable to tobacco smoking among adults in Ethiopia, from 1990 to 2023

**DOI:** 10.64898/2026.01.27.26345019

**Authors:** Sebsibe Tadesse, Gudina Egata, Mihretu Tagesse, Abraham Lomboro, Hiwot Abera, Yohannes Addisu Wondimagegne, Abraham Geremew, Sina Temesgen, Hirut Teame, Medhin Mehari, Hailu Lemma

## Abstract

**Background:** Non-communicable diseases are the result of a combination of genetic, physiological, environmental and behavioral factors. Evidence revealed that tobacco smoking is a leading cause of CVDs-related disability and premature mortality among others. Nevertheless, there is dearth of evidence on national and substantial health risks and distribution of health losses from CVDs associated with tobacco smoking in Ethiopia.

**Methods:** Data on mortality, disability-adjusted life years, years lived with disability, and years of life lost from CVDS attributable to tobacco smoking were extracted from the global burden of diseases 2023 study for Ethiopia and regions and administrative cities from 1990 to 2023. Comparative Risk Assessment Framework was used to generate the estimates. Disability-adjusted life years were obtained by adding the estimates of years lived with disability and years of life lost. Spatiotemporal Gausian process regression technique was employed to smooth the estimates. Rates were estimated per 100,000 population.

**Results:** An estimated 98332.1 (95% UI: 81623.8, 116279.3) CVDs-related mortalities occurred among adults aged 20 years and above in Ethiopia in 2023. The corresponding age-standardized death rate of CVDs attributable to tobacco smoking was estimated to be 221.1 deaths per 100,000 population (95% UI: 182.1, 261.5). A higher than the national age standardized death rate was estimated in Afar [266.2 (95% UI: 205.1, 332.2)], Benishangul-Gumuz [268.3 (95% UI: 216.9, 321.3)], South west [334.9 (95% UI: 250.3, 459.7)], and Addis Ababa [342.8 (95% UI: 260.6, 418.6)]. The age-standardized rate was estimated to be 5317.8 disability-adjusted life years (95% UI: 4503.3, 6237.6), 375.9 years lived with disability (95% UI: 275.9, 488.9), and 4941.9 years of life lost (95% UI: 4152.4, 5809.5). Stroke and ischemic heart disease were found to be the leading causes of deaths attributable to tobacco smoking. There was no significant trend shift in all the rates from 1990 to 2023.

**Conclusion:** This study has revealed that tobacco smoking continued to inflict substantial burden of disability and mortality among adults aged 20 years and above in Ethiopia, with subnational variation and stable trend over the past three decades. Ischemic stroke, ischemic heart disease, and intracerebral hemorrhage were found to be the leading causes of disability and premature mortality.

## Introduction

Non-communicable diseases (NCDs) are the result of a combination of genetic, physiological, environmental and behavioural factors [1]. Cardiovascular diseases (CVDs) being the main types of NCDs refer to a group of disorders of the heart and blood vessels [2]. In 2021, an estimated 19 million people died from NCDs mainly due to CVDs followed by others before the age of 70 years. Of which, 82% occurred in the low- and middle-income countries [1]. More than four out of five CVD deaths are due to heart attacks and strokes. Evidence revealed that tobacco smoking is a leading cause of CVD morbidity and mortality among others [3, 4].

The World Health Organization’s (WHO) global action plan on NCDs, outlines nine voluntary targets for the prevention and control of NCDs by the year 2025, which included a 25% relative reduction in risk of premature mortality from CVDs, cancer, diabetes, or chronic respiratory diseases, at least 10% relative reduction in the harmful use of alcohol, and a 30% relative reduction in prevalence of current tobacco use in persons aged 15 years and above [5]. In cognizant of the increase in burden of CVDs among adults, the government of Ethiopia also adapted different initiatives, such as development of stringent Alcohol and Framework Convention on Tobacco Control (FCTC) and strategic plan for control and prevention of major NCDs, including CVDs [6].

Despite the global and national efforts to reduce tobacco use, Ethiopia still faces substantial health risks associated with tobacco smoking, and yet limited evidence exists on the distribution of health losses from CVDs attributable to tobacco smoking at national and subnational levels. Thus, this study aims to bridge the knowledge gap by assessing the trends of mortality, years lived with disability (YLDs), years of life lost (YLLs), and disability-adjusted life years (DALYs) from CVDs attributable to tobacco smoking among adults aged 20 years and above by sex and locations in Ethiopia from 1990 to 2023, using data, methods, and tools of the Global Burden of Diseases (GBD) 2023 study. The study findings may provide evidence-based insights to design prevention and control strategies.

## Methods

### Study setting

This study was carried out in Ethiopia, a landlocked country located in the Horn of Africa, known for its diverse landscapes, rich history, and cultural heritage. Its recent population is estimated to be approximately 126.5 million in 2023, making it the 10^th^ most populous country globally and the second most populous in Africa after Nigeria. As of 2022, Ethiopia had a median age of 19.5 years, a fertility rate of 4.1 births per woman, and a life expectancy of 65 years [7, 8] Administratively, the country is divided into regional states, namely Oromia, Southern Nations, Nationalities, and Peoples (SNNP), Sidama, South West, Afar, Amhara, Somali, Tigray, Gambella, Benishangul-Gumuz, and Harari and city administrations at Addis Ababa and Dire Dawa.

### Data sources

The GBD 2023 study data were used to compute the mortality, YLDs, YLLs, and DALYs from CVDs attributable to tobacco smoking among adults aged 20 years and above at national and subnational levels in Ethiopia, from 1990 to 2023. The input data included health surveys, tobacco surveys, risk factors surveys, hospital records, and verbal autopsies. The full list of these data are available at: (https://ghdx.healthdata.org/gbd-2023/sources).

### Data analyses

For this analysis, the GBD 2023 study’s Comparative Risk Assessment Framework (CRAF) was used to estimate mortality and burden of CVDs attributable to tobacco smoking. The framework computed the theoretical minimum risk exposure level (TMREL), at which the lowest possible health losses from CVDs was observed [9]. The YLLs for each risk-outcome pair were the products of the total YLLs from CVDs attributable to tobacco smoking and the population-attributable fraction. The attributable-YLDs were calculated in the same way. The DALYs were estimated by adding YLLs and YLDs. The YLLs were calculated by multiplying the observed deaths by the reference age-specific life expectancy. The YLDs were determined by multiplying the prevalence of sequelae from CVDs attributable to tobacco smoking by the disability weight, which quantified the relative severity of the sequelae on a scale from 0 (perfect health) to 1 (death). Spatiotemporal Gausian process regression technique was employed to smooth the estimates. The presence of statistically significant trend changes was declared when the 95% uncertainty intervals (95% UIs) did not cross the zero value. To account for variations in data and modeling processes, all metrics were calculated with their 95% UIs. Rates were estimated per 100,000 populations.

## Results

### Mortality

An estimated 98332.1 (95% UI: 81623.8, 116279.3) CVDs-related mortalities occurred among adults aged 20 years and above in Ethiopia in 2023, with 51824.8 deaths among males (95% UI: 39876.3, 65717.4) and 46507.3 deaths among females (95% UI: (34535.7, 60886.2). Stroke and ischemic heart disease were found to be the leading causes of deaths attributable to tobacco smoking. The corresponding age-standardized death rate was estimated to be 221.1 deaths per 100,000 population (95% UI: 182.1, 261.5), with 225.1 deaths per 100,000 male population (95% UI: 174.1, 284.8) and 217.1 deaths per 100,000 female population (95% UI: 161.5, 282.8). A higher than the national age standardized death rate was estimated among Afar [266.2 (95% UI: 205.1, 332.2), Benishangul-Gumuz [268.3 (95% UI: 216.9, 321.3), South west [334.9 (95% UI: 250.3, 459.7), and Addis Ababa [342.8 (95% UI: 260.6, 418.6)] population. The high age standardized death rate was also observed among male population of these compared the national age standardized death rate of male population, 225.1 deaths per 100,000 populations (95% UI: 174.1, 284.8). From 1990 to 2023, there was no statistically significant trend shift in the mortality rate sub-nationally. There was an estimated 50 deaths per 100,000 population (95% UI: 40.0, 62.0) among adults aged 20 to 54 years, with 52 deaths per 100,000 males (95% UI: 36.8, 68.6) and 48 deaths per 100,000 females (95% UI: 33.0, 67.3). A higher than this rate was estimated among males in Benishangul-Gumuz [74.1 deaths per 100,000 males (95% UI: 48.8, 102.1)], South West [103.3 deaths per 100,000 males (95% UI: 61.6, 171.9)], and males in Addis Ababa [99.4 deaths per 100,000 males (95% UI: 66.7, 139.6)].

Nationally, there was no a significant increase in the death rate from 1990 to 2023. Similar estimates were observed in most of the regions in the country. However, the trend shift was not statistically significant among males and females in Tigray [a 0.6% (95% UI: (0.0, 1.7)], Amhara [a 0 % (95% UI: -0.4, 0.6)], Oromia [a -0.1%,(95% UI: -0.4, 0.5)], Somali [a -0.1%,(95% UI: -0.4, 0.5)],SNNPR [a 0.3% (95% UI: -0.3, 1.4)], Gambella [a 0.0% (95% UI: -0.4, 0.8)], Harari [a 0.0% ((95% UI: -0.3, 0.7)], DireDawa [a0.1% (95% UI: -0.3, 0.9)], Sidama [a 0.0% (95% UI: -0.5, 0.9)], South west [a 0.2% (95%UI: -0.3, 1.5)], Addis Ababa [a 0.0% (95%UI: (-0.3, 0.7)].and males in Afar [a 0.4% (95% UI: -0.2, 2.1))] and Benishangul-Gumuz[a 0.7% (95% UI: -0.1, 2.8)] except females in Benishangul-Gumuz [a 2.8% increase (95%UI: (0.9, 8.0)].

Among individuals aged 55 years and above, the rate was observed to be 1001.5 deaths per 100,000 population (95% UI: (823.9, 1182.8), with 1023.7 deaths per 100,000 males (95% UI: 801.5, 1297.9) and 977.4 deaths per 100,000 females (95% UI: 736.0, 1262.5). A higher than this estimate was observed among males in Addis Ababa [1774.4 deaths per 100,000 males (95% UI: 1333.8, 2169.5)]. There was no a significant increase in the death rate among individuals aged 55 years and above nationally and sub nationally from 1990 to 2023. Stroke [78 deaths per 100,000 population (95% UI: 55, 100)] and ischemic heart disease [56 deaths per 100,000 population (95% UI: 40, 80)] were found to be the leading causes of death attributable to tobacco smoking (Table 1, Figure 1).

**Table 1:** Mortalities from CVDs attributable to tobacco smoking among adults by sex, age, and location in Ethiopia, 1990 to 2023.

| Location | Sex | Number, age ≥20 years | Age-standardized rate | Annualized rate of change | Rate, age 20-54 years | Annualized rate of change | Rate, age ≥55 years | Annualized rate of change |
| --- | --- | --- | --- | --- | --- | --- | --- | --- |
|  |  | 2023 | 2023 | 1990 - 2023 | 2023 | 1990 - 2023 | 2023 | 1990-2023 |
| Ethiopia | Male | 51824.8 (39876.3, 65717.4) | 225.1 (174.1, 284.8) | 0.3 (-0.2, 1.5) | 51.6 (36.8, 68.6) | -0.2 (-0.5, 0.8) | 1023.7 (801.5, 1297.9) | 0.7 (0.1, 2.4) |
|  | Female | 46507.3 (34535.7, 60886.2) | 217.1 (161.5, 282.8) | 0.5 (-0.1, 2.2) | 47.5 (33.0, 67.3) | 0.3 (-0.3, 2.2) | 977.4 (736.0, 1262.5) | 0.6 (0.0, 2.4) |
|  | Both | 98332.1 (81623.8, 116279.3) | 221.1 (182.1, 261.5) | 0.4 (-0.1, 1.1) | 49.5 (40.0, 62.0) | 0.0 (-0.4, 0.7) | 1001.5 (823.9, 1182.8) | 0.7 (0.2, 1.6) |
| Tigray | Male | 3526.8 (2630.8, 4572.9) | 202.5 (151.5, 262.3) | 0.8 (0.1, 2.6) | 54.1 (36.7, 80.3) | 0.5 (-0.3, 2.7) | 996.5 (747.9, 1268.9) | 1.2 (0.3, 3.3) |
|  | Female | 3348.4 (2492.1, 4432.8) | 199.8 (146.3, 268.1) | 0.9 (0.1, 2.8) | 43.5 (27.3, 62.9) | 0.6 (-0.1, 2.7) | 980.1 (690.1, 1326.3) | 1.1 (0.2, 2.9) |
|  | Both | 6875.2 (5573.3, 8379.2) | 200.6 (162.5, 244.7) | 0.8 (0.2, 1.9) | 48.6 (36.2, 65.0) | 0.6 (0.0, 1.7) | 988.4 (792.6, 1194.4) | 1.2 (0.4, 2.5) |
| Afar | Male | 801.0 (514.8, 1064.9) | 266.7 (173.7, 352.2) | 0.8 (0.2, 2.5) | 57.2 (33.9, 78.5) | 0.4 (-0.2, 2.1) | 1141.7 (745.2, 1530.1) | 1.3 (0.5, 3.4) |
|  | Female | 455.7 (331.1, 614.4) | 268.5 (196.6, 354.9) | 0.5 (-0.2, 1.8) | 50.6 (32.8, 75.9) | 1.8 (0.4, 5.2) | 996.9 (724.3, 1319.5) | 0.6 (-0.1, 2.3) |
|  | Both | 1256.8 (961.7, 1567.0) | 266.2 (205.1, 332.2) | 0.7 (0.2, 1.5) | 54.0 (39.1, 72.4) | 0.8 (0.2, 2.2) | 1090.3 (839.0, 1348.5) | 1.0 (0.5, 1.9) |
| Amhara | Male | 14543.1 (10913.4, 18985.7) | 231.4 (174.8, 302.6) | 0.2 (-0.2, 1.5) | 51.9 (36.6, 70.4) | -0.3 (-0.6, 0.5) | 1094.3 (830.9, 1421.2) | 0.7 (0.1, 2.4) |
|  | Female | 13897.7 (10311.9, 17968.7) | 235.3 (173.2, 306.8) | 0.8 (0.1, 2.9) | 50.7 (34.9, 71.3) | 0.5 (-0.2, 2.7) | 1106.6 (816.7, 1443.9) | 0.9 (0.1, 2.9) |
|  | Both | 28440.8 (23186.3, 33926.7) | 233.1 (188.5, 277.6) | 0.5 (0.0, 1.3) | 51.3 (40.1, 65.1) | 0.0 (-0.4, 0.6) | 1100.2 (885.5, 1311.6) | 0.8 (0.3, 1.7) |
| Oromia | Male | 15333.8 (11718.5, 19588.1) | 211.7 (162.5, 272.3) | 0.2 (-0.3, 1.5) | 43.0 (29.6, 59.2) | -0.2 (-0.6, 0.8) | 968.6 (743.4, 1246.7) | 0.9 (0.2, 2.9) |
|  | Female | 13908.6 (10062.4, 18393.3) | 197.9 (139.6, 258.3) | 0.2 (-0.3, 1.8) | 41.9 (28.3, 60.3) | 0.1 (-0.4, 1.6) | 885.1 (627.6, 1180.0) | 0.6 (-0.1, 2.7) |
|  | Both | 29242.4 (23802.1, 35059.8) | 204.8 (165.7, 245.2) | 0.2 (-0.2, 0.9) | 42.4 (33.1, 54.9) | -0.1 (-0.4, 0.5) | 927.7 (748.3, 1114.5) | 0.7 (0.1, 1.8) |
| Somali | Male | 2916.6 (2074.9, 3835.7) | 203.9 (147.9, 272.3) | 0.3 (-0.2, 1.2) | 50.0 (31.9, 70.9) | -0.4 (-0.6, 0.1) | 867.1 (621.8, 1140.0) | 0.3 (-0.2, 1.1) |
|  | Female | 1344.2 (983.4, 1862.7) | 167.2 (121.2, 228.5) | 0.1 (-0.4, 0.8) | 41.0 (28.0, 61.7) | 0.8 (0.0, 2.6) | 618.9 (448.5, 849.2) | 0.0 (-0.4, 0.7) |
|  | Both | 4260.8 (3381.1, 5298.0) | 191.5 (147.7, 240.3) | 0.2 (-0.1, 0.8) | 45.7 (33.7, 61.4) | -0.1 (-0.4, 0.5) | 782.3 (603.5, 973.0) | 0.2 (-0.1, 0.9) |
| Benishangul-Gumuz | Male | 774.1 (588.7, 991.5) | 293.7 (224.1, 375.4) | 0.3 (-0.2, 1.6) | 74.1 (48.8, 102.1) | 0.7 (-0.1, 2.8) | 1238.7 (951.7, 1555.8) | 0.8 (0.1, 2.5) |
|  | Female | 507.9 (385.5, 679.0) | 238.0 (178.3, 314.9) | 0.4 (-0.2, 1.8) | 53.2 (36.5, 79.7) | 2.8 (0.9, 8.0) | 950.9 (719.3, 1229.7) | 0.6 (-0.1, 2.3) |
|  | Both | 1282.0 (1025.5, 1583.8) | 268.3 (216.9, 321.3) | 0.3 (-0.1, 1.0) | 63.8 (47.0, 83.3) | 1.2 (0.3, 3.1) | 1108.7 (904.3, 1309.2) | 0.7 (0.3, 1.6) |
| SNNP | Male | 5944.5 (4370.1, 8434.8) | 183.9 (134.8, 259.4) | 0.4 (-0.2, 2.4) | 49.2 (33.6, 74.4) | 0.1 (-0.4, 2.0) | 777.4 (572.0, 1093.0) | 0.8 (0.0, 3.2) |
|  | Female | 6906.4 (4754.1, 9487.3) | 213.1 (146.2, 290.3) | 0.6 (-0.1, 2.9) | 54.6 (34.4, 83.5) | 0.4 (-0.3, 3.1) | 911.5 (620.6, 1239.7) | 0.5 (-0.2, 2.5) |
|  | Both | 12850.9 (10157.3, 16129.9) | 199.1 (155.9, 254.3) | 0.5 (-0.1, 1.6) | 52.0 (39.1, 70.9) | 0.3 (-0.3, 1.4) | 843.4 (653.5, 1075.3) | 0.7 (0.0, 1.8) |
| Gambella | Male | 186.9 (134.0, 262.1) | 243.8 (175.9, 332.5) | 0.0 (-0.4, 1.1) | 47.9 (31.4, 71.7) | 0.1 (-0.4, 1.8) | 946.1 (681.2, 1283.4) | 0.3 (-0.3, 1.8) |
|  | Female | 173.9 (120.3, 234.7) | 224.1 (158.7, 304.5) | -0.5 (-0.7, 0.0) | 45.1 (28.1, 65.0) | -0.1 (-0.6, 1.0) | 715.0 (503.4, 969.5) | -0.4 (-0.7, 0.2) |
|  | Both | 360.8 (284.0, 452.5) | 232.9 (185.7, 289.5) | -0.4 (-0.6, 0.1) | 46.4 (31.1, 61.9) | 0.0 (-0.4, 0.8) | 822.7 (660.6, 1028.6) | -0.2 (-0.5, 0.4) |
| Harari | Male | 120.4 (90.3, 155.3) | 228.1 (171.9, 292.0) | 1.5 (0.5, 4.5) | 55.3 (36.0, 78.3) | 0.3 (-0.2, 2.1) | 945.8 (720.5, 1211.4) | 1.6 (0.6, 4.4) |
|  | Female | 140.5 (101.8, 190.2) | 239.2 (173.7, 319.3) | -0.1 (-0.4, 0.7) | 58.9 (37.3, 81.4) | -0.2 (-0.5, 0.7) | 1026.2 (757.6, 1370.6) | -0.1 (-0.4, 0.7) |
|  | Both | 260.9 (207.1, 322.4) | 234.4 (187.2, 290.3) | 0.2 (-0.1, 1.0) | 57.0 (40.6, 74.3) | 0.0 (-0.3, 0.7) | 988.7 (788.3, 1208.4) | 0.1 (-0.2, 0.8) |
| Dire Dawa | Male | 245.5 (175.8, 330.5) | 227.3 (162.2, 306.5) | 0.4 (-0.2, 2.0) | 58.2 (38.4, 87.3) | 0.1 (-0.5, 1.6) | 935.5 (670.3, 1242.6) | 0.3 (-0.2, 1.6) |
|  | Female | 243.5 (176.8, 325.3) | 214.2 (154.0, 286.9) | 0.0 (-0.4, 1.0) | 48.4 (31.8, 72.3) | 0.1 (-0.4, 1.5) | 875.6 (627.6, 1178.8) | -0.1 (-0.5, 0.7) |
|  | Both | 489.1 (385.2, 603.3) | 220.5 (173.7, 269.2) | 0.2 (-0.2, 0.9) | 53.3 (37.8, 72.2) | 0.1 (-0.3, 0.9) | 903.7 (712.1, 1103.1) | 0.0 (-0.3, 0.6) |
| Sidama | Male | 1771.4 (1144.3, 2801.4) | 201.9 (130.9, 316.6) | 0.0 (-0.5, 1.4) | 44.4 (25.6, 78.4) | -0.2 (-0.7, 1.1) | 955.3 (629.1, 1480.2) | 0.3 (-0.3, 2.0) |
|  | Female | 1555.5 (1031.1, 2483.0) | 206.4 (137.2, 323.9) | 0.2 (-0.4, 1.8) | 46.0 (28.0, 82.8) | 0.3 (-0.4, 2.8) | 941.2 (626.9, 1471.8) | 0.2 (-0.3, 1.7) |
|  | Both | 3326.8 (2454.1, 4490.8) | 203.9 (150.9, 273.3) | 0.1 (-0.3, 0.9) | 45.2 (31.2, 65.4) | 0.0 (-0.5, 0.9) | 948.9 (698.1, 1262.7) | 0.3 (-0.2, 1.2) |
| South West | Male | 1931.1 (1305.4, 2969.2) | 388.8 (264.3, 600.1) | 0.3 (-0.3, 1.9) | 103.3 (61.6, 171.9) | 0.2 (-0.5, 2.2) | 1723.8 (1181.2, 2649.4) | 0.6 (-0.1, 2.5) |
|  | Female | 1141.6 (736.4, 1846.0) | 273.1 (175.5, 432.4) | 0.3 (-0.3, 2.1) | 63.4 (36.3, 113.9) | 0.3 (-0.4, 3.1) | 1142.8 (735.2, 1802.7) | 0.6 (-0.1, 2.7) |
|  | Both | 3072.7 (2300.3, 4265.9) | 334.9 (250.3, 459.7) | 0.3 (-0.2, 1.3) | 82.8 (56.8, 118.9) | 0.2 (-0.3, 1.5) | 1455.9 (1090.7, 1983.8) | 0.6 (0.0, 1.8) |
| Addis Ababa | Male | 3729.4 (2769.4, 4616.5) | 403.8 (303.7, 491.6) | 0.6 (0.1, 1.7) | 99.4 (66.7, 139.6) | 0.0 (-0.4, 0.9) | 1774.4 (1333.8, 2169.5) | 0.6 (0.1, 1.5) |
|  | Female | 2883.5 (1618.6, 3862.6) | 288.6 (156.4, 381.3) | 0.6 (0.1, 1.5) | 56.0 (30.5, 86.8) | 0.1 (-0.4, 1.2) | 1311.4 (713.3, 1733.8) | 0.4 (0.0, 1.0) |
|  | Both | 6612.9 (5054.6, 8155.7) | 342.8 (260.6, 418.6) | 0.6 (0.2, 1.3) | 76.3 (56.5, 102.1) | 0.0 (-0.3, 0.7) | 1531.1 (1155.5, 1856.2) | 0.5 (0.1, 1.0) |

**Figure 1.**
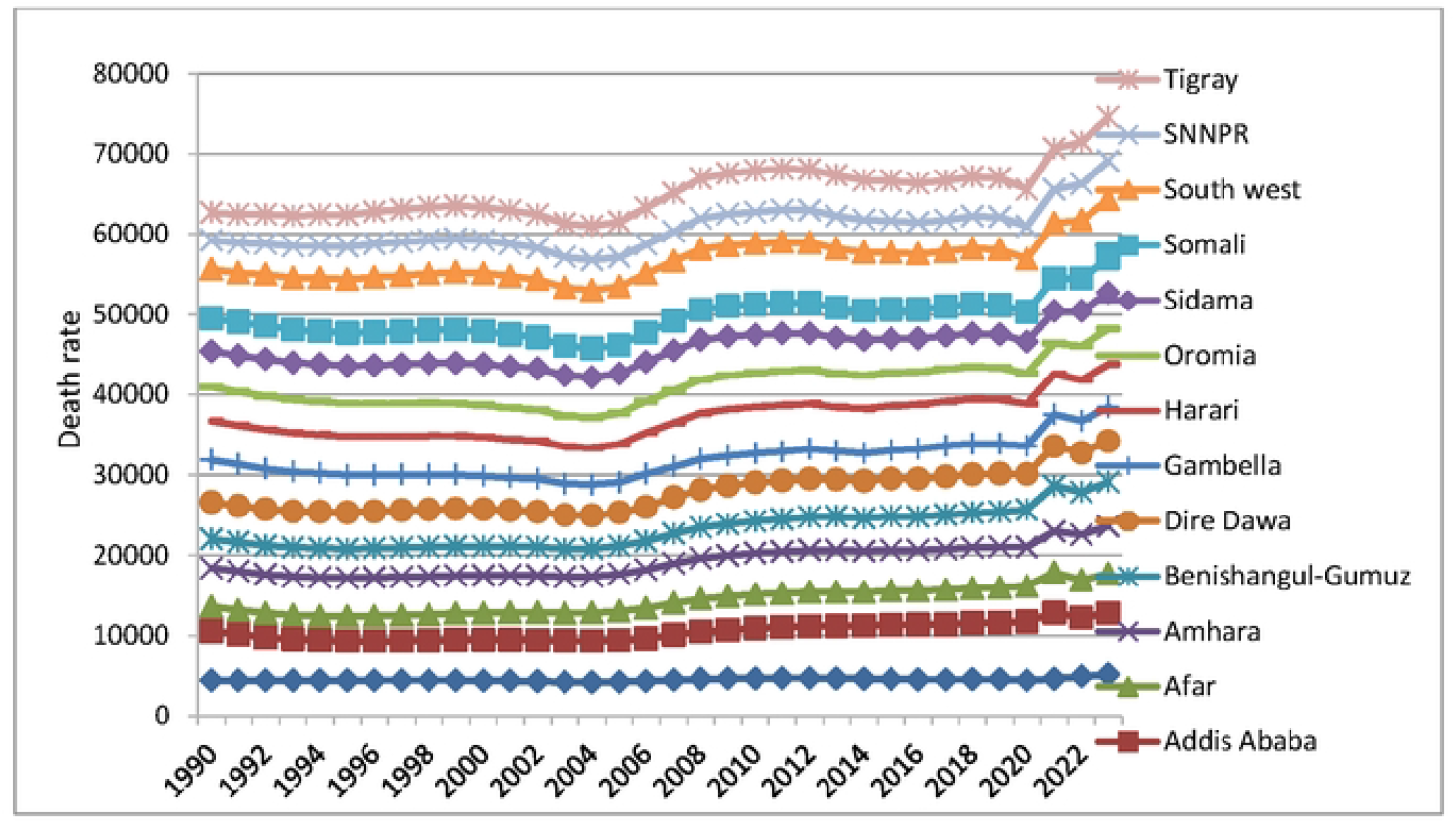
Death rates from CVDs attributable to tobacco smoking among adults aged 20 years and above in Ethiopia, 1990 to 2023

### Disability-adjusted life years

The age-standardized rate of DALYs from CVDs attributable to tobacco smoking was estimated to be 5317.8 years per 100,000 population (95% UI: 4503.3, 6237.6) among adults aged 20 years and above in Ethiopia in 2023 with 5515.8 DALYs per 100,000 males (95% UI: 4349.0, 6951.1) and 5113.3 DALYs per 100,000 females (95% UI: 3932.3, 6623.9). A higher than this rate was estimated among males in Benishangul-Gumuz [7300.7 DALYs per 100,000 males (95% UI: 5693.3, 9188.8), South West [9670.9 DALYs per 100,000 males (95% UI: 6682.2, 14715.4), and males in Addis Ababa [9286.4 DALYs per 100,000 males (95% UI: (6997.3, 11305.8)]. There was no significant increase in the DALYs rate among both sexes from 1990 to 2023. Nationally, the rate of DALYs increased as age increased (Table 2, Figure 2, Figure 3).

**Table 2:**
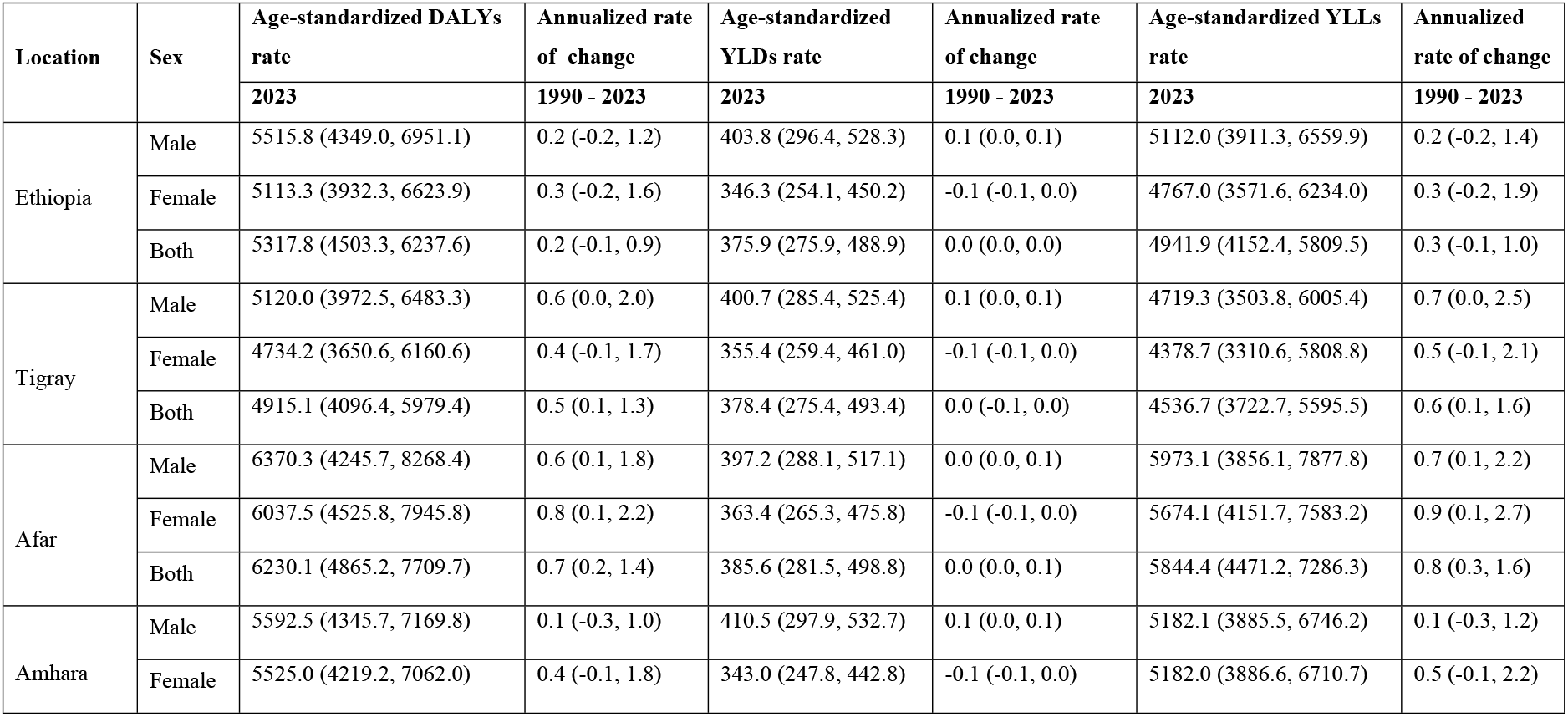
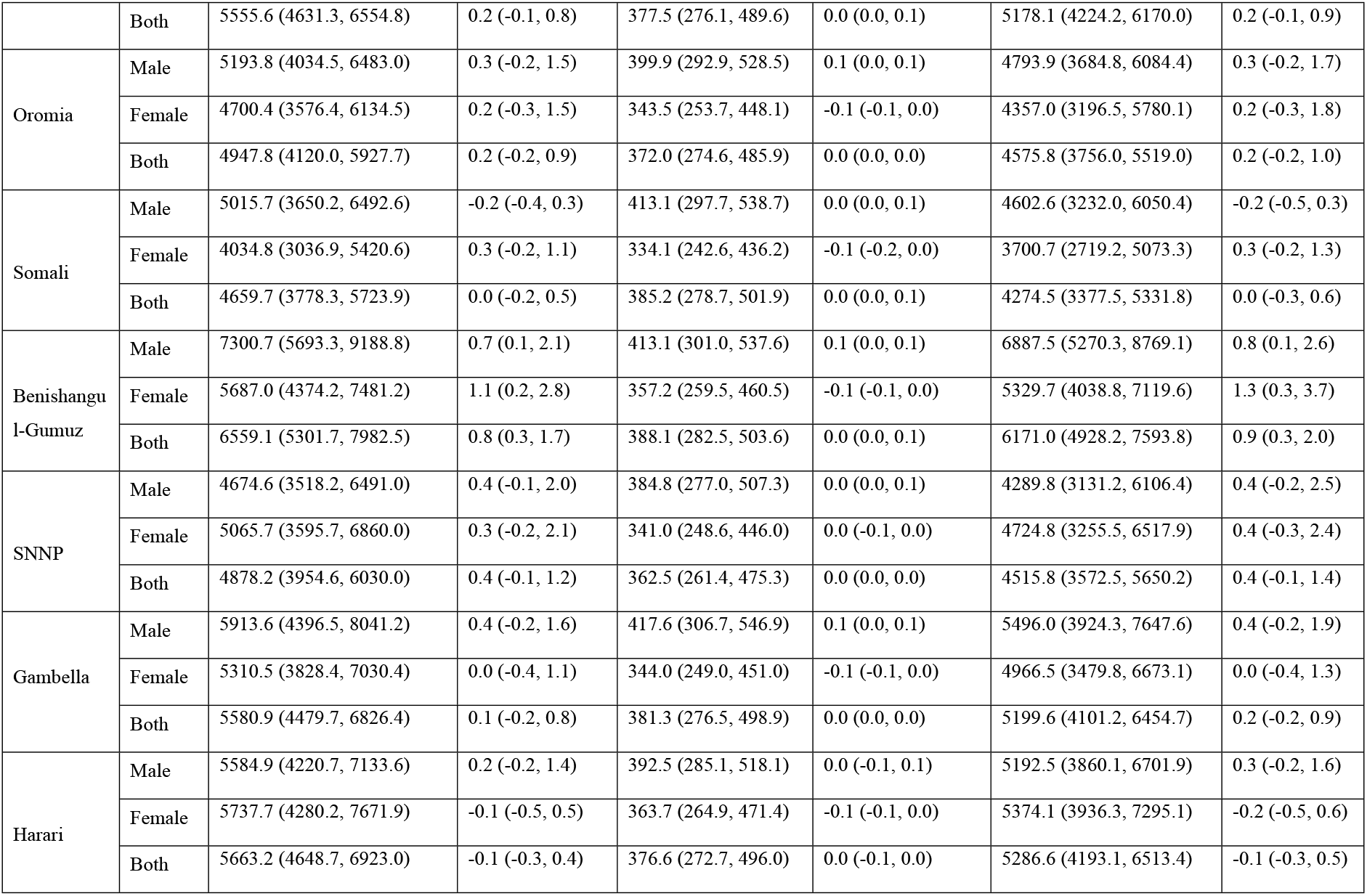
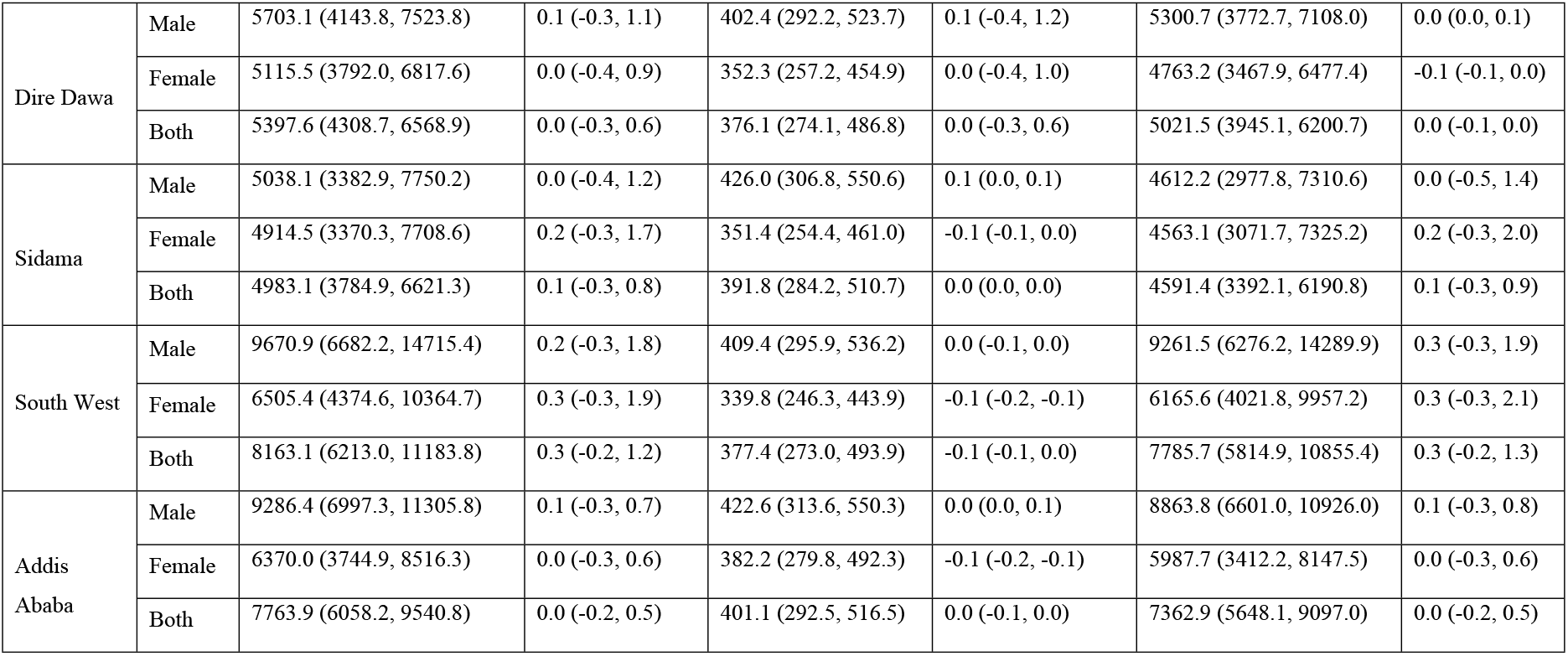
Age-standardized rates of DALYs, YLDs, and YLLs, by sex and location in Ethiopia, 1990 to 2023.

| Location | Sex | Age-standardized DALYs rate | Annualized rate of change | Age-standardized YLDs rate | Annualized rate of change | Age-standardized YLLs rate | Annualized rate of change |
| --- | --- | --- | --- | --- | --- | --- | --- |
|  |  | 2023 | 1990 - 2023 | 2023 | 1990 - 2023 | 2023 | 1990 - 2023 |
| Ethiopia | Male | 5515.8 (4349.0, 6951.1) | 0.2 (-0.2, 1.2) | 403.8 (296.4, 528.3) | 0.1 (0.0, 0.1) | 5112.0 (3911.3, 6559.9) | 0.2 (-0.2, 1.4) |
|  | Female | 5113.3 (3932.3, 6623.9) | 0.3 (-0.2, 1.6) | 346.3 (254.1, 450.2) | -0.1 (-0.1, 0.0) | 4767.0 (3571.6, 6234.0) | 0.3 (-0.2, 1.9) |
|  | Both | 5317.8 (4503.3, 6237.6) | 0.2 (-0.1, 0.9) | 375.9 (275.9, 488.9) | 0.0 (0.0, 0.0) | 4941.9 (4152.4, 5809.5) | 0.3 (-0.1, 1.0) |
| Tigray | Male | 5120.0 (3972.5, 6483.3) | 0.6 (0.0, 2.0) | 400.7 (285.4, 525.4) | 0.1 (0.0, 0.1) | 4719.3 (3503.8, 6005.4) | 0.7 (0.0, 2.5) |
|  | Female | 4734.2 (3650.6, 6160.6) | 0.4 (-0.1, 1.7) | 355.4 (259.4, 461.0) | -0.1 (-0.1, 0.0) | 4378.7 (3310.6, 5808.8) | 0.5 (-0.1, 2.1) |
|  | Both | 4915.1 (4096.4, 5979.4) | 0.5 (0.1, 1.3) | 378.4 (275.4, 493.4) | 0.0 (-0.1, 0.0) | 4536.7 (3722.7, 5595.5) | 0.6 (0.1, 1.6) |
| Afar | Male | 6370.3 (4245.7, 8268.4) | 0.6 (0.1, 1.8) | 397.2 (288.1, 517.1) | 0.0 (0.0, 0.1) | 5973.1 (3856.1, 7877.8) | 0.7 (0.1, 2.2) |
|  | Female | 6037.5 (4525.8, 7945.8) | 0.8 (0.1, 2.2) | 363.4 (265.3, 475.8) | -0.1 (-0.1, 0.0) | 5674.1 (4151.7, 7583.2) | 0.9 (0.1, 2.7) |
|  | Both | 6230.1 (4865.2, 7709.7) | 0.7 (0.2, 1.4) | 385.6 (281.5, 498.8) | 0.0 (0.0, 0.1) | 5844.4 (4471.2, 7286.3) | 0.8 (0.3, 1.6) |
| Amhara | Male | 5592.5 (4345.7, 7169.8) | 0.1 (-0.3, 1.0) | 410.5 (297.9, 532.7) | 0.1 (0.0, 0.1) | 5182.1 (3885.5, 6746.2) | 0.1 (-0.3, 1.2) |
|  | Female | 5525.0 (4219.2, 7062.0) | 0.4 (-0.1, 1.8) | 343.0 (247.8, 442.8) | -0.1 (-0.1, 0.0) | 5182.0 (3886.6, 6710.7) | 0.5 (-0.1, 2.2) |
|  | Both | 5555.6 (4631.3, 6554.8) | 0.2 (-0.1, 0.8) | 377.5 (276.1, 489.6) | 0.0 (0.0, 0.1) | 5178.1 (4224.2, 6170.0) | 0.2 (-0.1, 0.9) |
| Oromia | Male | 5193.8 (4034.5, 6483.0) | 0.3 (-0.2, 1.5) | 399.9 (292.9, 528.5) | 0.1 (0.0, 0.1) | 4793.9 (3684.8, 6084.4) | 0.3 (-0.2, 1.7) |
|  | Female | 4700.4 (3576.4, 6134.5) | 0.2 (-0.3, 1.5) | 343.5 (253.7, 448.1) | -0.1 (-0.1, 0.0) | 4357.0 (3196.5, 5780.1) | 0.2 (-0.3, 1.8) |
|  | Both | 4947.8 (4120.0, 5927.7) | 0.2 (-0.2, 0.9) | 372.0 (274.6, 485.9) | 0.0 (0.0, 0.0) | 4575.8 (3756.0, 5519.0) | 0.2 (-0.2, 1.0) |
| Somali | Male | 5015.7 (3650.2, 6492.6) | -0.2 (-0.4, 0.3) | 413.1 (297.7, 538.7) | 0.0 (0.0, 0.1) | 4602.6 (3232.0, 6050.4) | -0.2 (-0.5, 0.3) |
|  | Female | 4034.8 (3036.9, 5420.6) | 0.3 (-0.2, 1.1) | 334.1 (242.6, 436.2) | -0.1 (-0.2, 0.0) | 3700.7 (2719.2, 5073.3) | 0.3 (-0.2, 1.3) |
|  | Both | 4659.7 (3778.3, 5723.9) | 0.0 (-0.2, 0.5) | 385.2 (278.7, 501.9) | 0.0 (0.0, 0.1) | 4274.5 (3377.5, 5331.8) | 0.0 (-0.3, 0.6) |
| Benishangu<br>I-Gumuz | Male | 7300.7 (5693.3, 9188.8) | 0.7 (0.1, 2.1) | 413.1 (301.0, 537.6) | 0.1 (0.0, 0.1) | 6887.5 (5270.3, 8769.1) | 0.8 (0.1, 2.6) |
|  | Female | 5687.0 (4374.2, 7481.2) | 1.1 (0.2, 2.8) | 357.2 (259.5, 460.5) | -0.1 (-0.1, 0.0) | 5329.7 (4038.8, 7119.6) | 1.3 (0.3, 3.7) |
|  | Both | 6559.1 (5301.7, 7982.5) | 0.8 (0.3, 1.7) | 388.1 (282.5, 503.6) | 0.0 (0.0, 0.1) | 6171.0 (4928.2, 7593.8) | 0.9 (0.3, 2.0) |
| SNNP | Male | 4674.6 (3518.2, 6491.0) | 0.4 (-0.1, 2.0) | 384.8 (277.0, 507.3) | 0.0 (0.0, 0.1) | 4289.8 (3131.2, 6106.4) | 0.4 (-0.2, 2.5) |
|  | Female | 5065.7 (3595.7, 6860.0) | 0.3 (-0.2, 2.1) | 341.0 (248.6, 446.0) | 0.0 (-0.1, 0.0) | 4724.8 (3255.5, 6517.9) | 0.4 (-0.3, 2.4) |
|  | Both | 4878.2 (3954.6, 6030.0) | 0.4 (-0.1, 1.2) | 362.5 (261.4, 475.3) | 0.0 (0.0, 0.0) | 4515.8 (3572.5, 5650.2) | 0.4 (-0.1, 1.4) |
| Gambella | Male | 5913.6 (4396.5, 8041.2) | 0.4 (-0.2, 1.6) | 417.6 (306.7, 546.9) | 0.1 (0.0, 0.1) | 5496.0 (3924.3, 7647.6) | 0.4 (-0.2, 1.9) |
|  | Female | 5310.5 (3828.4, 7030.4) | 0.0 (-0.4, 1.1) | 344.0 (249.0, 451.0) | -0.1 (-0.1, 0.0) | 4966.5 (3479.8, 6673.1) | 0.0 (-0.4, 1.3) |
|  | Both | 5580.9 (4479.7, 6826.4) | 0.1 (-0.2, 0.8) | 381.3 (276.5, 498.9) | 0.0 (0.0, 0.0) | 5199.6 (4101.2, 6454.7) | 0.2 (-0.2, 0.9) |
| Harari | Male | 5584.9 (4220.7, 7133.6) | 0.2 (-0.2, 1.4) | 392.5 (285.1, 518.1) | 0.0 (-0.1, 0.1) | 5192.5 (3860.1, 6701.9) | 0.3 (-0.2, 1.6) |
|  | Female | 5737.7 (4280.2, 7671.9) | -0.1 (-0.5, 0.5) | 363.7 (264.9, 471.4) | -0.1 (-0.1, 0.0) | 5374.1 (3936.3, 7295.1) | -0.2 (-0.5, 0.6) |
|  | Both | 5663.2 (4648.7, 6923.0) | -0.1 (-0.3, 0.4) | 376.6 (272.7, 496.0) | 0.0 (-0.1, 0.0) | 5286.6 (4193.1, 6513.4) | -0.1 (-0.3, 0.5) |
| Dire Dawa | Male | 5703.1 (4143.8, 7523.8) | 0.1 (-0.3, 1.1) | 402.4 (292.2, 523.7) | 0.1 (-0.4, 1.2) | 5300.7 (3772.7, 7108.0) | 0.0 (0.0, 0.1) |
|  | Female | 5115.5 (3792.0, 6817.6) | 0.0 (-0.4, 0.9) | 352.3 (257.2, 454.9) | 0.0 (-0.4, 1.0) | 4763.2 (3467.9, 6477.4) | -0.1 (-0.1, 0.0) |
|  | Both | 5397.6 (4308.7, 6568.9) | 0.0 (-0.3, 0.6) | 376.1 (274.1, 486.8) | 0.0 (-0.3, 0.6) | 5021.5 (3945.1, 6200.7) | 0.0 (-0.1, 0.0) |
| Sidama | Male | 5038.1 (3382.9, 7750.2) | 0.0 (-0.4, 1.2) | 426.0 (306.8, 550.6) | 0.1 (0.0, 0.1) | 4612.2 (2977.8, 7310.6) | 0.0 (-0.5, 1.4) |
|  | Female | 4914.5 (3370.3, 7708.6) | 0.2 (-0.3, 1.7) | 351.4 (254.4, 461.0) | -0.1 (-0.1, 0.0) | 4563.1 (3071.7, 7325.2) | 0.2 (-0.3, 2.0) |
|  | Both | 4983.1 (3784.9, 6621.3) | 0.1 (-0.3, 0.8) | 391.8 (284.2, 510.7) | 0.0 (0.0, 0.0) | 4591.4 (3392.1, 6190.8) | 0.1 (-0.3, 0.9) |
| South West | Male | 9670.9 (6682.2, 14715.4) | 0.2 (-0.3, 1.8) | 409.4 (295.9, 536.2) | 0.0 (-0.1, 0.0) | 9261.5 (6276.2, 14289.9) | 0.3 (-0.3, 1.9) |
|  | Female | 6505.4 (4374.6, 10364.7) | 0.3 (-0.3, 1.9) | 339.8 (246.3, 443.9) | -0.1 (-0.2, -0.1) | 6165.6 (4021.8, 9957.2) | 0.3 (-0.3, 2.1) |
|  | Both | 8163.1 (6213.0, 11183.8) | 0.3 (-0.2, 1.2) | 377.4 (273.0, 493.9) | -0.1 (-0.1, 0.0) | 7785.7 (5814.9, 10855.4) | 0.3 (-0.2, 1.3) |
| Addis Ababa | Male | 9286.4 (6997.3, 11305.8) | 0.1 (-0.3, 0.7) | 422.6 (313.6, 550.3) | 0.0 (0.0, 0.1) | 8863.8 (6601.0, 10926.0) | 0.1 (-0.3, 0.8) |
|  | Female | 6370.0 (3744.9, 8516.3) | 0.0 (-0.3, 0.6) | 382.2 (279.8, 492.3) | -0.1 (-0.2, -0.1) | 5987.7 (3412.2, 8147.5) | 0.0 (-0.3, 0.6) |
|  | Both | 7763.9 (6058.2, 9540.8) | 0.0 (-0.2, 0.5) | 401.1 (292.5, 516.5) | 0.0 (-0.1, 0.0) | 7362.9 (5648.1, 9097.0) | 0.0 (-0.2, 0.5) |

**Figure 2.**
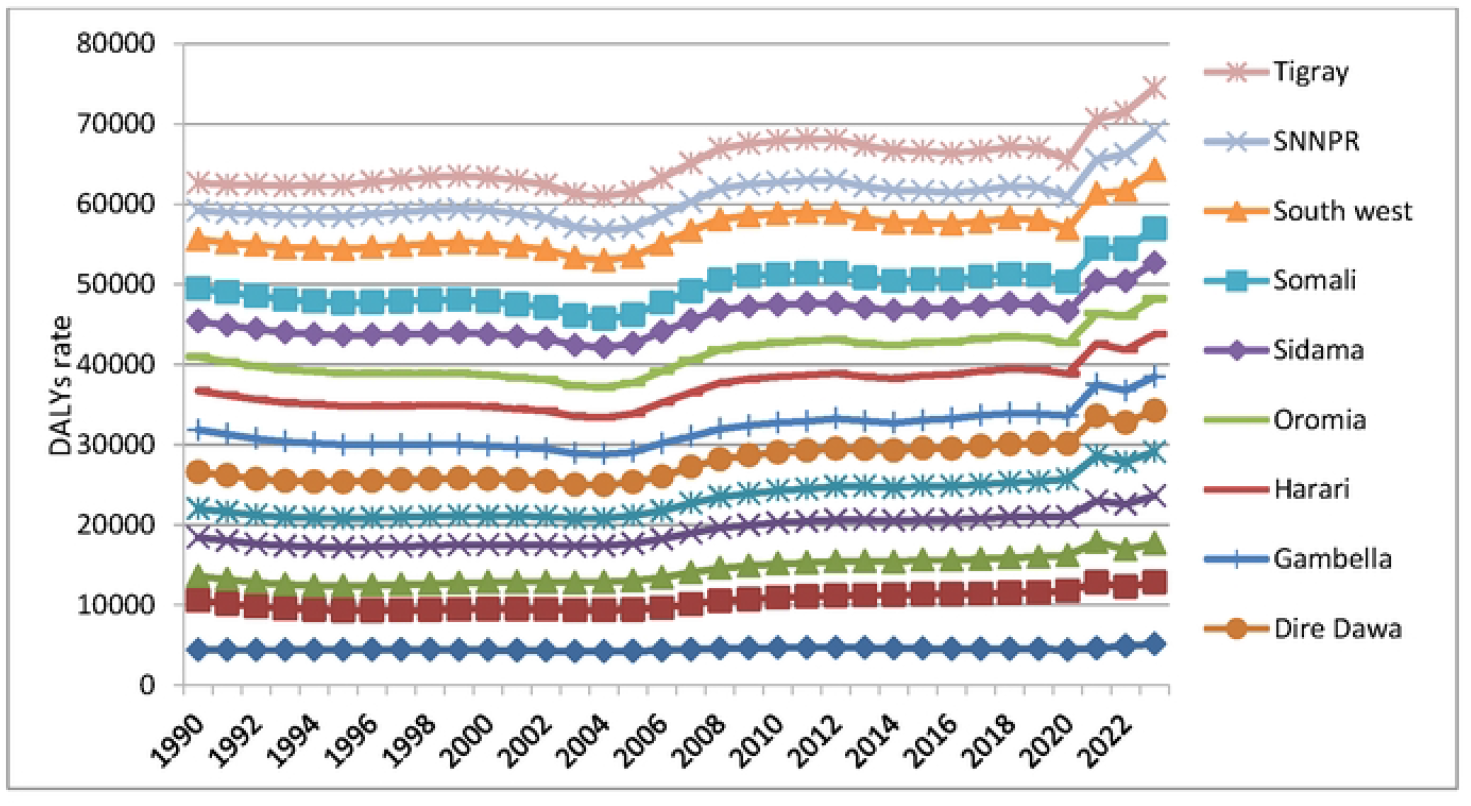
DALYs rates from CVDs attributable to tobacco smoking among adults aged 20years and above in Ethiopia, 1990 to 2023

**Figure 3.**
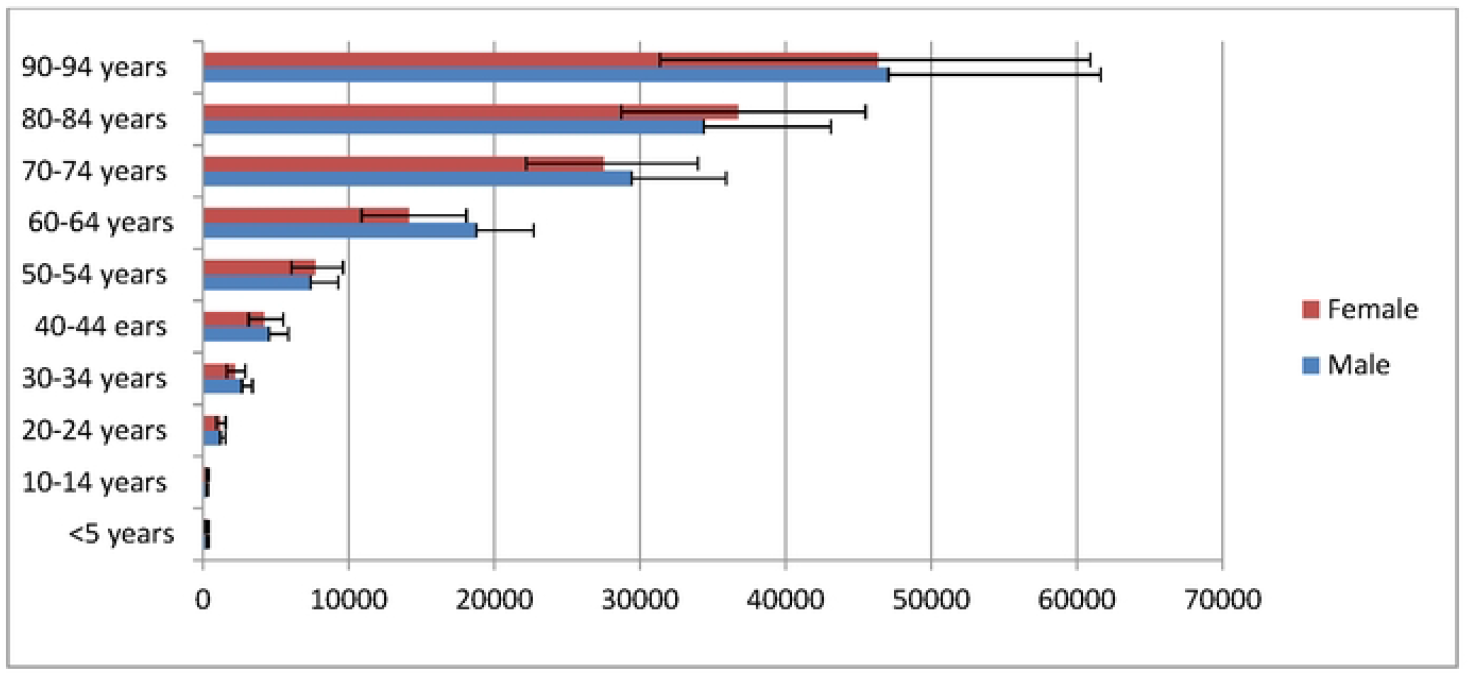
Age and sex distribution of DALYs rate from CVDs attributable to tobacco smoking in Ethiopia, 2023

### Years lived with disability

The age-standardized rate of YLDs from CVDs attributable to tobacco smoking was estimated to be 375.9 years per 100,000 population (95% UI: 275.9, 488.9) among adults aged 20 years and above in Ethiopia in 2023, with 403.8 YLDS per 100,000 males (95% UI: 296.4, 528.3) and 346.3 YLDs per 100,000 females (95% UI: 254.1, 450.2). There was no significant increase in the YLDs rate among both sexes from 1990 to 2023 (Table 2). However, ischemic stroke, ischemic heart disease, and intracerebral hemorrhage were found to the leading causes of YLDs among adults aged 20 years and above in Ethiopia in 2023 (Table 2, Figure 4).

**Figure 4.**
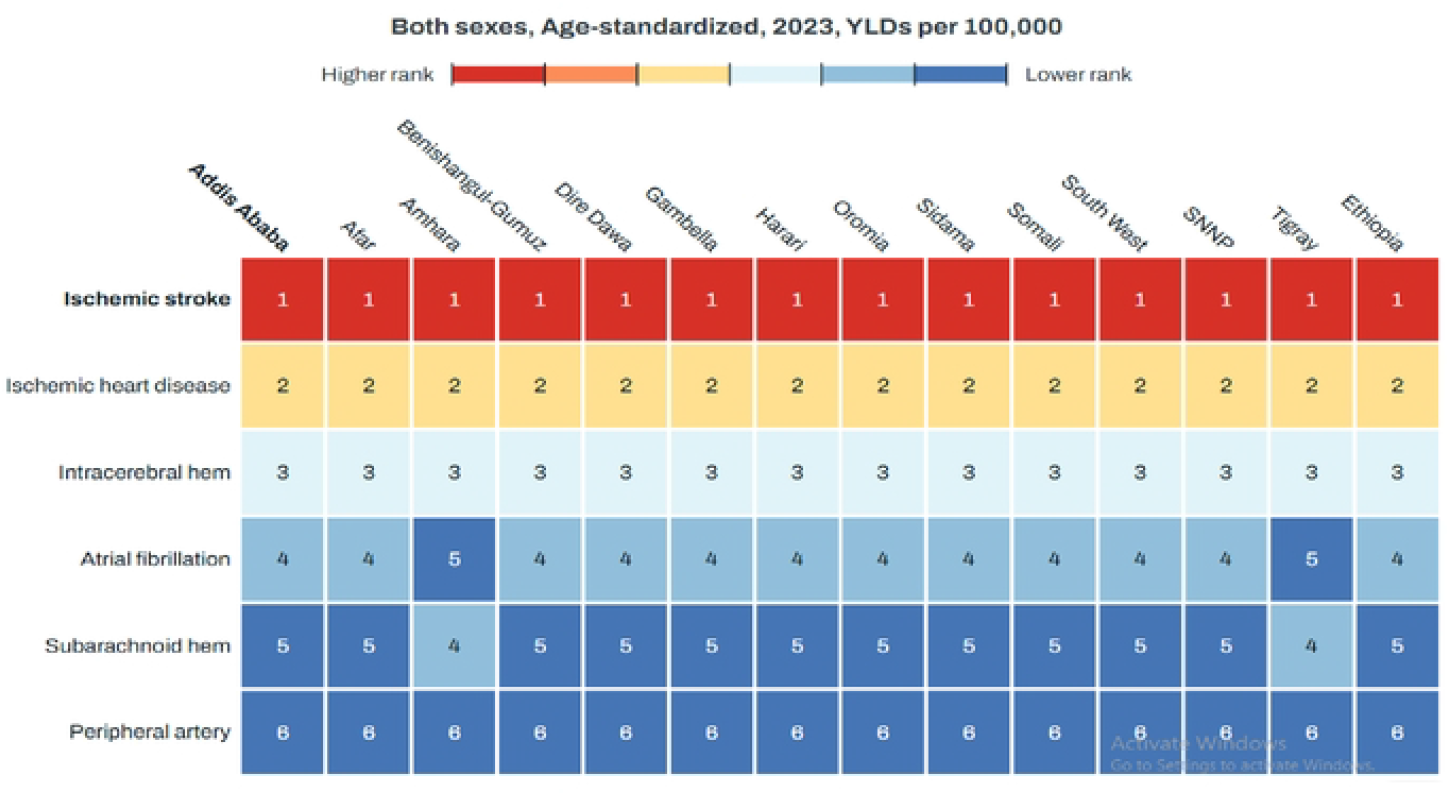
Age-standardized YLDs rates from the leading CVDs attributable to tobacco smoking in Ethiopia, 2023

### Years of life lost

The age-standardized rate of YLLs from CVDs attributable to tobacco smoking was estimated to be 4941.9 years per 100,000 population (95% UI: 4152.4, 5809.5) among adults aged 20 years and above in Ethiopia in 2023, with 5112.0 YLLs per 100,000 males (95% UI: (3911.3, 6559.9) and 4767.0YLLs per 100,000 females (95% UI: (3571.6, 6234.0). A higher than this rate was estimated among males in Benishangul-Gumuz [6887.5 YLLs per 100,000 males (95% UI: 5270.3, 8769.1)], males in South West [9261.5 YLLs per 100,000 males (95% UI: (6276.2, 14289.9)], and Addis Ababa [8863.8 YLLs per 100,000 males (95% UI: (6601.0, 10926.0)]. There was no significant increase in the YLLs rate among both sexes from 1990 to 2023. Ischemic heart disease, intracerebral hemorrhage, and ischemic stroke were the leading CVDs attributable to tobacco smoking, causing higher age-standardized YLLs rates nationally and subnationally (Table 2, Figure 5, Figure 6).

**Figure 5.**
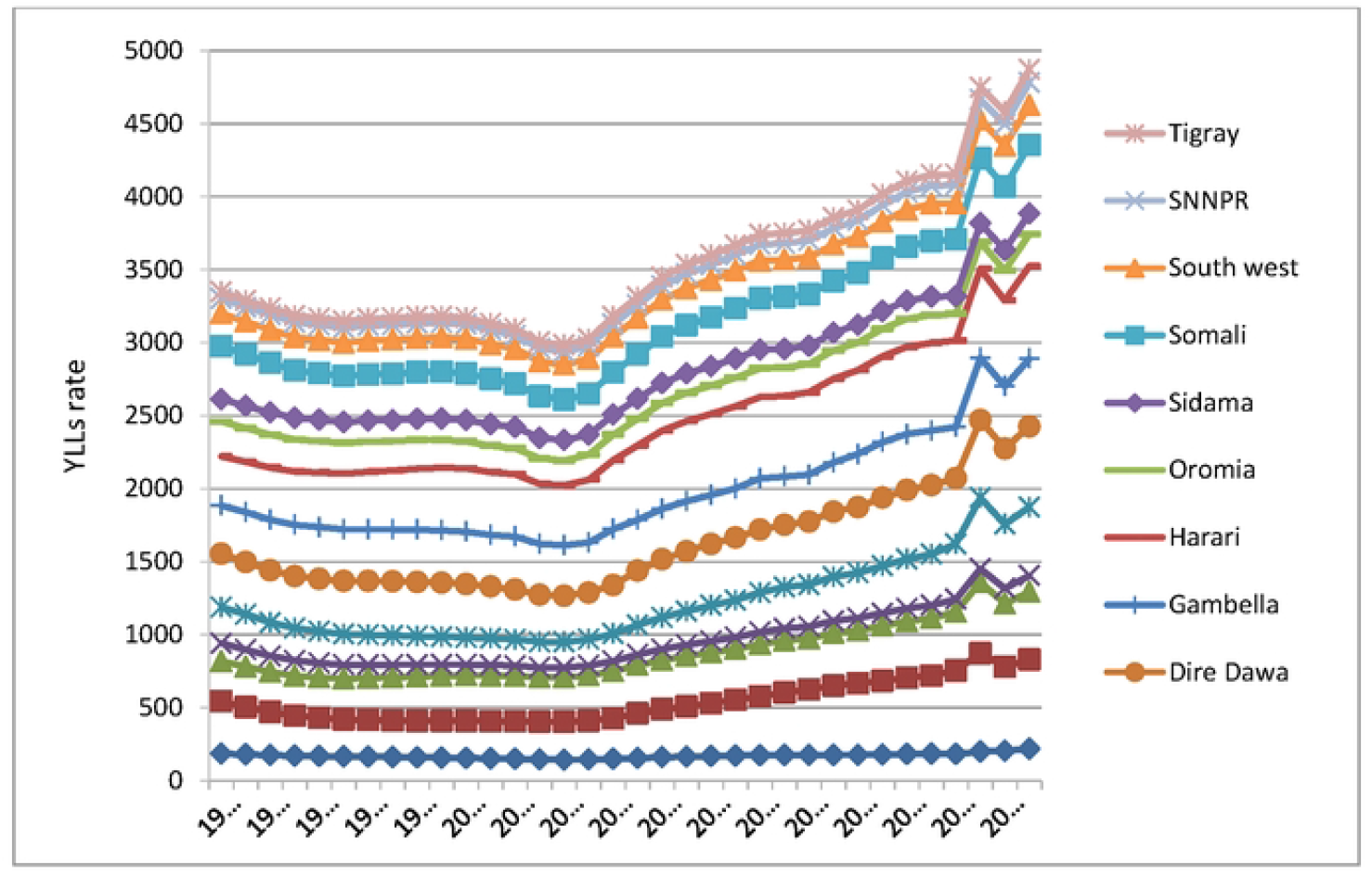
YLLs rates from CVDs attributable to tobacco smoking among adults aged 20years and above in Ethiopia, 1993 to 2023

**Figure 6.**
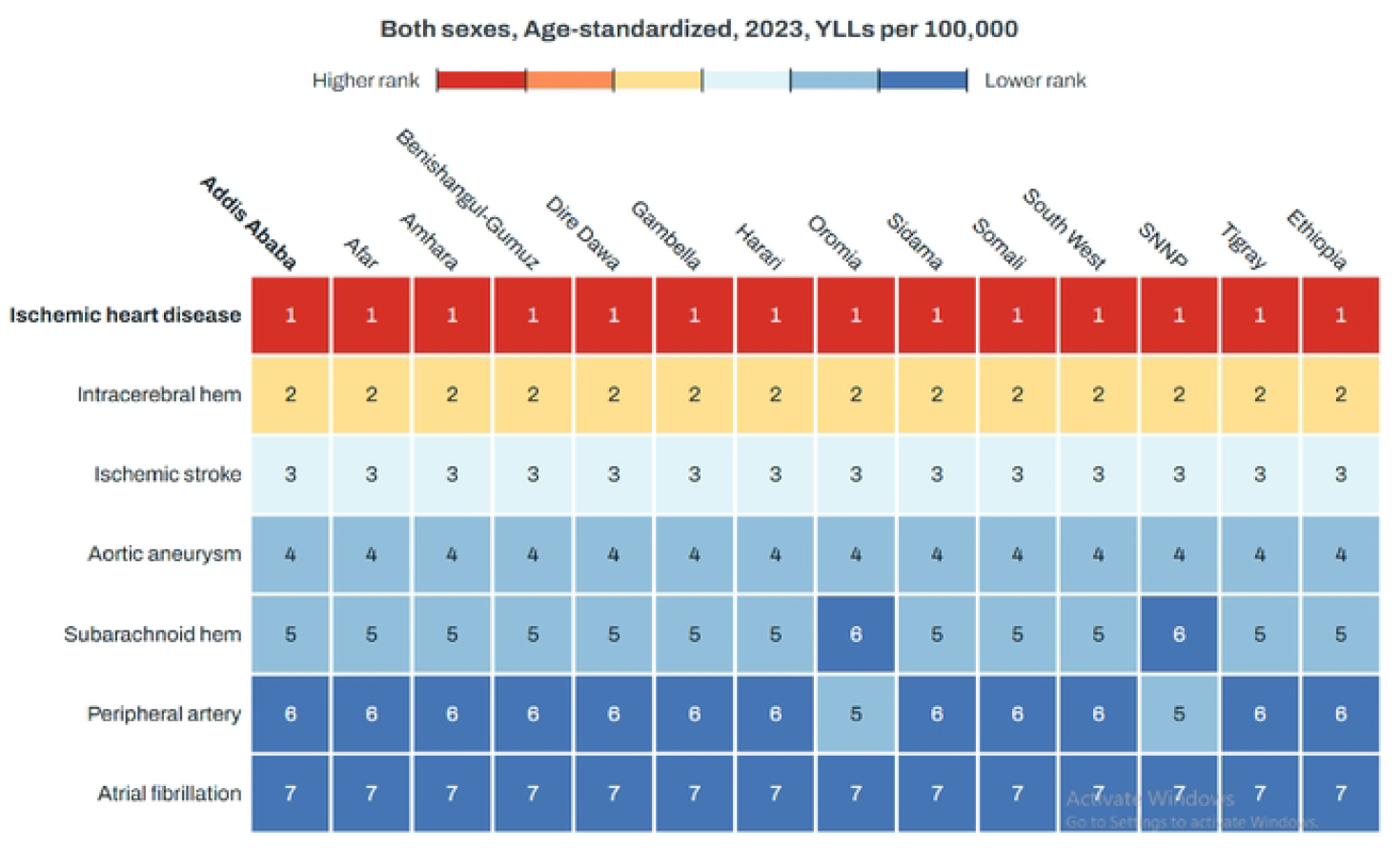
Age-standardized YLLs rates from the leading CVDs attributable to tobacco smoking in Ethiopia, 2023

## Discussion

The present analysis was undertaken to estimate the national and sub–national burden and trend of CVDs attributed to tobacco smoking in Ethiopia from 1990 to 2023 using the GBD study data and methods. Accordingly, an estimated 98,332.1 CVDs-related mortalities (95% UI: 81623.8, 116279.3) occurred among adults aged 20 years and above in Ethiopia in 2023. The corresponding age-standardized rate of mortality was 221.1 deaths per 100,000 population (95% UI: 182.1, 261.5), DALYs was 5317.8 years per 100,000 population (95% UI: 4503.3, 6237.6), YLDs was 375.9 years per 100,000 population (95% UI: 275.9, 488.9), and YLLs was 4941.9 years per 100,000 population (95% UI: 4152.4, 5809.5). Ischemic heart disease, intracerebral hemorrhage, and ischemic stroke were the leading causes of age-standardized rates of YLDs and YLLs from CVDs attributable to tobacco smoking nationally and sub-nationally.

An estimated 98,332.1 deaths from CVDs attributable to tobacco smoking (95% UI: 81623.8, 116279.3) occurred among adults aged 20 years and above in Ethiopia in 2023. This was lower than the global number of deaths from CVDs attributable to smoking which has increased from 1.8 million in 1990 to 2.2 million in 2021, representing a 26.16% rise. This figure was also lower than that of East Asia which was reported as 0.8 million deaths but greater than that of Oceania, which was reported to be 3,456 [10]. There is no variation in mortality for males and females in this study unlike another study which has evaluated cardiovascular risk of smoking and reported that the mortality from CVDs is higher in female than male smokers [11].

On the other hand, the corresponding age-standardized CVDs related mortality rate attributed to tobacco smoking was 221.1 deaths per 100,000 population (95% UI: 182.1, 261.5) in this analysis. This was consistent with the finding of the GBD 2017 which has reported age-standardized CVD mortality in Ethiopia as 182.63 deaths per 100,000 population (95% UI: 165.49 – 203.9). However, although the authors analyze risk-attribution, including risk factors for CVD, they did not break out a mortality rate specifically attributed to tobacco use [12]. Specifically, there was high age standardized CVDs mortality rate attributed to tobacco use among population from Afar, Benishangul-Gumuz, South West, and Addis Ababa. Afar region had also experienced the highest age-standardized all-cause death rate which was estimated to be 1353.38 deaths per 100 000 population (95% UI:1195·69, 1526·19)[13]. The high age-standardized death rate was also higher among male population of these regions compared to the national age standardized death rate of male population. This high age-standardized mortality rate estimated among males is consistent with the GBD 2021 study results, which indicated consistently higher burden of smoking-related CVDs in males [10].

The age-standardized rate of DALYs from CVDs attributable to tobacco smoking was estimated to be 5317.8 years per 100,000 population (95% UI: 4503.3, 6237.6) among adults aged 20 years and above in Ethiopia in 2023. The observed DALYs rate of the GBD 2023 estimate is greater than the result of GBD 2017 study in 1990 which was 3549.6 DALYs per 100 000 population (95% UI : 3229.0 - 3911.9) in 2017 [12] but is lower than the result of GBD 2021 for men were smoking at the global level [124.1 million DALYs (95% UI 111.2 million to 137.0 million)] [9]. The DALYs estimate was more common among individuals aged 20-24 years with males aged 60-64 years and above are more affected compared to males of lower age boundaries. The commonness of DALYs among males is in accordance with the result of 2017 GBD study [12]. Cumulatively, ischemic stroke, ischemic heart disease, and intracerebral hemorrhage were found to be the leading causes of YLDs and YLLs among adults aged 20 years and above in Ethiopia in 2023.This finding is in congruence with 1990–2019: a subnational country analysis for the Global Burden of Disease Study 2019 when stroke, and ischaemic heart disease were identified as the leading causes of premature mortality alongside other non-communicable disease for all sexes in Ethiopia in 2019 [13].

With regard to trend shift in mortality rate from CVDs attributed to tobacco smoking, there was no statistically significant trend shift sub-nationally from 1990 to 2023. However, there was a slight increase in Tigray, Afar, and Addis Ababa. Likewise, there was slight increase in the rate of DALYs and YLLs among males and females in Afar and Benishangul-Gumuz from 1990 to 2023. Nevertheless, there was no significant increase in the YLDs rate among both sexes in the same years. Since there was no exactly similar subnational analysis done before, it is difficult to compare our result. The age-standardized all-cause death rate was the highest in Afar region in 2021 [1353 per 100 000 population (95% UI: 1195·69–1526·19)] [13]. In this study, Afar region had the highest mortality rate from CVDs attributed to tobacco smoking [221deaths per 100,000 populations (95% UI: 182, 262] in the country in 2023.

This study could have the following limitations: Mortality and smoking prevalence data at subnational level in Ethiopia may be incomplete, inconsistent, or missing for some years and regions, which can bias estimates or increase uncertainty. Vital registration and verbal autopsy systems can misclassify cardiovascular deaths (e.g., stroke versus other causes), leading to error in attributing deaths to CVDs and to tobacco-attributable fractions. Smoking exposure is often self-reported and may under-report prevalence, intensity, or duration; subnational survey coverage may be uneven, producing measurement error in exposure inputs. GBD-style models typically rely on population-level associations; they may not account for subnational differences in confounders that modify tobacco–CVDs relationships. The study may apply relative risks derived from international cohorts to Ethiopian subpopulations and these relative risks may not reflect local genetics, comorbidities, or healthcare access, biasing estimates. Subnational estimates often rely on statistical smoothing or covariate-based models where direct data are sparse; model structure and covariate choice can strongly influence local estimates. Inaccurate or outdated population estimates and internal migration can bias rate calculations and trends at subnational levels. Although uncertainty intervals may be reported, wide intervals for regions and years with sparse data reduce precision and complicate interpretation of trends and differences. Estimates may not accurately represent marginalized or small subpopulations (e.g., displaced persons, remote rural communities) that are underrepresented in surveys.

Overall, the findings of this study highlight that tobacco-attributable cardiovascular mortality and health losses remain a major and persistent public health challenge in Ethiopia, signaling the need for stronger and more targeted policy action. To reduce this burden, Ethiopia must accelerate enforcement of existing tobacco-control laws, increase tobacco taxation, expand smoke-free public spaces, and strengthen regulatory measures against illicit tobacco trade (14, 15). In practice, the health system should integrate routine screening for smoking in all clinical encounters, scale up accessible cessation services, and intensify public education campaigns that clearly communicate the cardiovascular risks of tobacco use. Prioritizing interventions in subnational regions with the highest burden will ensure more efficient allocation of resources and help achieve meaningful reductions in preventable deaths and disability.

## Conclusion

In sum, stroke and ischemic heart disease were found to be the leading causes of deaths attributable to tobacco smoking.The age standardized CVDs death rate attributable to tobacco smoking is substantial among adults aged 20 years and above in Ethiopia with some regions having high rate including Addis Ababa. Likewise, the age-standardized rate of DALYs rate from CVDs attributable to tobacco smoking was high among the same population with gender disparity for few regions. The premature mortality is nearly 5000 years per 100,000 populations with some gender disparity between few regions. Ischemic stroke, ischemic heart disease, and intracerebral hemorrhage were found to the leading causes of YLDs and premature mortality in these populations in 2023 both nationally and sub-nationally. Although there was no significant increase in the YLDs rate among both sexes, there was slight increase in the DALYs and YLLs per 100,000 population for both sexes in Afar and Benishangul-Gumuz.

## Data Availability

Data can be obtained from global burden of disease data base

## Data availability statement

The datasets presented in this article are not readily available because one can access them from the IHME repository directly. Requests to access the datasets should be directed to http://ghdx.647healthdata.org/gbd-results-tool

## Ethics statement

This study was adhered to the GBD protocol, and conducted as part of the GBD Collaborators Network (https://www.healthdata.org/research-analysis/about-gbd/protocol).

## Author contributions

ST conceived and designed the study. ST, GE, MT, AL, HA, YAW, AG, ST, HT, MM, and HL were involved in the analysis and interpretation of the findings. All authors have approved the final version of the manuscript.

## Acknowledgments

The GBD study is a systematic, scientific, and global effort to quantify and compare the magnitude of health loss from diseases, injuries, and risk factors by age, sex, and population over time. We are grateful for this global initiative.

## Conflict of interest

The authors declare that the research was conducted in the absence of any commercial or financial relationships that could be construed as a potential conflict of interest

## Notes

### Competing Interest Statement

The authors have declared no competing interest.

### Funding Statement

No fund was obtained for this manuscript writing

### Author Declarations

Ethical standards were followed

## References

1. World Health Oorganization. Key facts on non-communicable diseases. World Health Oorganization; 2024. Available at: https://www.who.int/news-room/fact-sheets/detail/noncommunicable-diseases. Date accessed: 26/Apr./2025.

2. World Health Organization. Key facts on cardiovascular diseases. World Health Oorganization; 2021. Available at: https://www.who.int/health-topics/cardiovascular-diseases#tab=tab_1. Date accessed: 25/Apr.2025. 2021.

3. Banks E, Joshy G, Korda RJ, Stavreski B, Soga K, Egger S, et al. Tobacco smoking and risk of 36 cardiovascular disease subtypes: fatal and non-fatal outcomes in a large prospective Australian study. BMC Medicine 2019; 17:128.

4. Joshy G, et al. Relationship of tobacco smoking to cause-specific mortality: contemporary estimates from Australi. 2025; 23:115.

5. World Health Organization. Global action plan for the prevention and control of non-communicable diseases 2013–2020. World Health Organization; 2013. Available at: https://iris.who.int/bitstream/handle/10665/94384/9789241506236_eng.pdf?sequence=1. Date accessed: 26/Apr./2025.

6. Federal Ministry of Health. National strategic plan for the prevention and control of major non-communicable diseases. Federal Ministry of Health; 2020. Available at: https://www.iccp-portal.org/system/files/plans/ETH_B3_s21_National_Strategic_Plan_for_Prevention_and_Control_of_NCDs2021.pdf. Date accessed: 26/Apr./2025.

7. World Bank. Ethiopia overview. World Bank, 2024. Available at: https://www.worldbank.org/en/country/ethiopia/overview. Date accessed: 03/July/2025.

8. United Nations. Wolrd population prospects 2024: Summary of results. Department of Economic and Social Affairs: United Nations, 2022. Available at: https://population.un.org/wpp/. Date accessed: 03/July/2023.

9. Stanaway JD, Afshin A, Gakidou E, Lim SS, Abate D, Abate KH, et al. Global, regional, and national comparative risk assessment of 84 behavioral, environmental and occupational, and metabolic risks or clusters of risks, 1990–2016: a systematic analysis for the Global Burden of Disease Study 2016. The Lancet 2017; 390:1345–1422..

10. Zhu S, Gao J, Zhang L, Dong W, Shi W, Guo H, et al. Global, regional, and national cardiovascular disease burden attributable to smoking from 1990 to 2021: Findings from the GBD 2021 Study. Tob Induc Dis 2025; 23:11.

11. Gallucci G, Tartarone A, Lerose R, Lalinga AV, Capobianco AM. Cardiovascular risk of smoking and benefits of smoking cessation. J Thorac Dis 2020; 12(7):3866–3876.

12. Ali S, Misganaw A, Worku A, Destaw Z, Negash L, Bekele A,, et al. The burden of cardiovascular diseases in Ethiopia from 1990 to 2017:evidence from the Global Burden of Disease Study. International Health 2021; 13: 318–326.

13. Misganaw A, Naghavi M, Walker A, Mirkuzie AH, Giref AZ, Berheto TM, et al. Progress in health among regions of Ethiopia, 1990–2019: a subnational country analysis for the Global Burden of Disease Study 2019. Lancet 2022; 399: 1322– 35.

14. Mamudu HM, Subedi P, Alamin AE, Veeranki SP, Owusu D, et al. The progress of tobacco control research in sub-saharan africa in the past 50 years: a systematic review of the design and methods of the studies. Int J Environ Res Public Health 2018; 15(12):2732

15. Reitsma MB, Kendrick PJ, Ababneh E, Abbafati C, Abbasi-Kangevari M, Abdoli A, et al: Spatial, temporal, and demographic patterns in prevalence of smoking tobacco use and attributable disease burden in 204 countries and territories, 1990-2019: a systematic analysis from the Global Burden of Disease Study 2019. The Lancet 2021, 397(10292):2337–2360.

